# Cardiovascular risk scores for primary prevention: head-to-head validation of 16 established and contemporary models

**DOI:** 10.64898/2026.07.02.26357120

**Authors:** Yuanning Hu, Shiqing Hu, Zilin Dong, Jiahui Wei, Zhongjie Zhang, Pengfei Jiang, Hao Huang, Tuo Li, Jian Zou

## Abstract

**Background and Aims:** Cardiovascular risk scores guide primary prevention, but their comparative performance remains uncertain. We externally validated 16 established and contemporary cardiovascular risk-prediction models in a common primary-prevention evaluation framework.

**Methods:** UK Biobank participants free from cardiovascular disease and cancer at baseline were included. 16 models, corresponding to 19 configurations, were implemented as published and evaluated against a harmonized incident CVD endpoint. Performance at 5 and 10 years was assessed using discrimination, calibration, Brier score, decision curve analysis, and TRIPOD+AI reporting quality.

**Results:** Among 438,640 participants, 45,003 incident cardiovascular events occurred over a median follow-up of 13.5 years. Ten-year area under the curve ranged from 0.668 for QRISK3 to 0.734 for PREDICT, and C-index from 0.655 to 0.717. Calibration varied substantially: PROCAM, Framingham, ASSIGN, and QRISK1 overestimated risk, whereas PREVENT, PREDICT, SCORE, SCORE2, SCORE2-OP, and China-PAR generally underestimated risk. QRISK2 showed the best calibration, while PREVENT had the lowest Brier score. At higher treatment thresholds, net benefit diverged, with PREVENT and PCE performing most consistently. Composite assessment favored PREVENT, QRISK2, PREDICT, PCE, and northern China-PAR variants.

**Conclusions:** Direct application of cardiovascular risk scores across populations can produce clinically important differences in calibration and net benefit. Model selection for primary prevention should require external validation, local recalibration, and assessment of clinical utility, rather than reliance on discrimination alone. PREVENT, QRISK2, PREDICT, PCE, and northern China-PAR variants showed the most balanced performance in this cohort.

## Introduction

Cardiovascular disease (CVD) remains one of the leading causes of death and disability worldwide, accounting for an estimated 20.5 million deaths in 2021, approximately one-third of all global deaths^1^. Since many effective preventive interventions are most useful before clinical disease becomes apparent, primary prevention increasingly depends on estimating an individual’s absolute future cardiovascular risk. Contemporary guidelines embed risk prediction into routine decision-making: the National Institute for Health and Care Excellence (NICE) recommends QRISK3 to estimate 10-year CVD risk in adults aged 25–84 years without established CVD^2^, whereas ACC/AHA guidance classifies adults into low, borderline, intermediate, and high 10-year ASCVD risk categories using the Pooled Cohort Equations (PCE)^3^. In Europe, SCORE2 and SCORE2-OP were introduced to estimate 10-year fatal and non-fatal CVD risk in middle-aged and older adults, respectively^4,5^, and have been incorporated into ESC cardiovascular prevention guidance and clinical risk-estimation tools^6^.

This proliferation of risk scores reflects a central challenge in cardiovascular prevention: risk models are not biologically universal instruments, but statistical products of the populations, eras, outcome definitions, predictors, and healthcare systems from which they were derived. Older models such as Framingham and PROCAM^7,8^ were developed in historical cohorts with higher baseline event rates and narrower population diversity. Later models, including QRISK^9^, PCE^10^, China-PAR^11^, PREDICT^12^, Reynolds^13,14^, SCORE2, WHO-CVD risk^15^, Globorisk^16^, and PREVENT^17^, attempted to address different limitations by incorporating contemporary data, regional recalibration, ethnicity, deprivation, kidney or metabolic markers, or global risk-region structures. SCORE2, for example, was derived from 45 European cohorts and recalibrated to four European risk regions; PREVENT was developed from contemporary data from more than 6.5 million diverse US adults and incorporates cardiovascular-kidney-metabolic factors. These methodological differences make direct comparison necessary before adopting or transferring a score to a new clinical setting.

External validation of CVD risk scores has been undertaken extensively but unevenly^18^. A systematic review by Damen *et al.* identified 363 prediction models for CVD in the general population and found that only a minority had been externally validated, with head-to-head comparisons of more than a few models in the same cohort exceptionally rare^19^. Subsequent UK Biobank validations have examined QRISK3 alone^20^ and PREVENT versus the PCE^21,22^, while large multi-cohort studies have compared up to four algorithms after systematic recalibration^23^. However, to the best of our knowledge, no study has simultaneously validated the full set of widely used or guideline-endorsed scores in a single contemporary cohort under harmonized analytical conditions. As a consequence, clinicians and guideline committees choosing between scores must rely on disparate validation reports that differ in cohort, outcome definition, follow-up length, statistical method, and the dimensions of performance evaluated. Critically, most validations report discrimination, fewer report calibration in a quantitative form suitable for cross-model comparison, and very few report clinical utility through decision curve analysis or assess the transparency of the original development study against contemporary reporting standards such as TRIPOD+AI^24,25^. A multi-domain, head-to-head comparison is therefore needed to inform rational model selection.

In this study, we externally validated 16 established and contemporary cardiovascular risk-prediction models, represented by 19 configurations, in 438,640 UK Biobank participants without prevalent cardiovascular disease or cancer at baseline. **Our primary interest was cardiovascular primary prevention: determining which scores most effectively identify initially disease-free adults who may benefit from preventive intervention before a first cardiovascular event.** Over up to 15 years of follow-up, during which 45,003 incident cardiovascular events occurred, we compared models under a harmonized evaluation framework that included discrimination, calibration, prediction error, decision-curve utility, and reporting transparency. We also conducted an exploratory multidomain assessment^26^ to examine whether these dimensions supported a consistent ranking or instead highlighted trade-offs relevant to model selection. These results provide a practical evidence base for choosing, recalibrating, and applying cardiovascular risk scores in primary-prevention populations.

## Methods

### Study design and data source

This validation study used data from 502,129 participants included the UK Biobank^27^, a large-scale prospective population-based cohort. All analyses were conducted under UK Biobank Application ID 160957. Ethical approval for the UK Biobank was granted by the North West Multi-Centre Research Ethics Committee (reference 11/NW/0382), and all participants provided written informed consent.

We constructed a primary-prevention cohort of adults without clinically manifest cardiovascular disease at baseline to compare cardiovascular risk-prediction models in a common population-level setting. Participants with prevalent cardiovascular disease (ICD-10: I20–I25, I60–I64, I46) were excluded to focus on first incident cardiovascular events. Participants with any malignant cancer except non-melanoma skin cancer (C44) were also excluded to reduce potential distortion from cancer-related mortality, treatment exposures, and prognosis-driven follow-up. A study flowchart is shown in Figure 1.

**Fig. 1:**
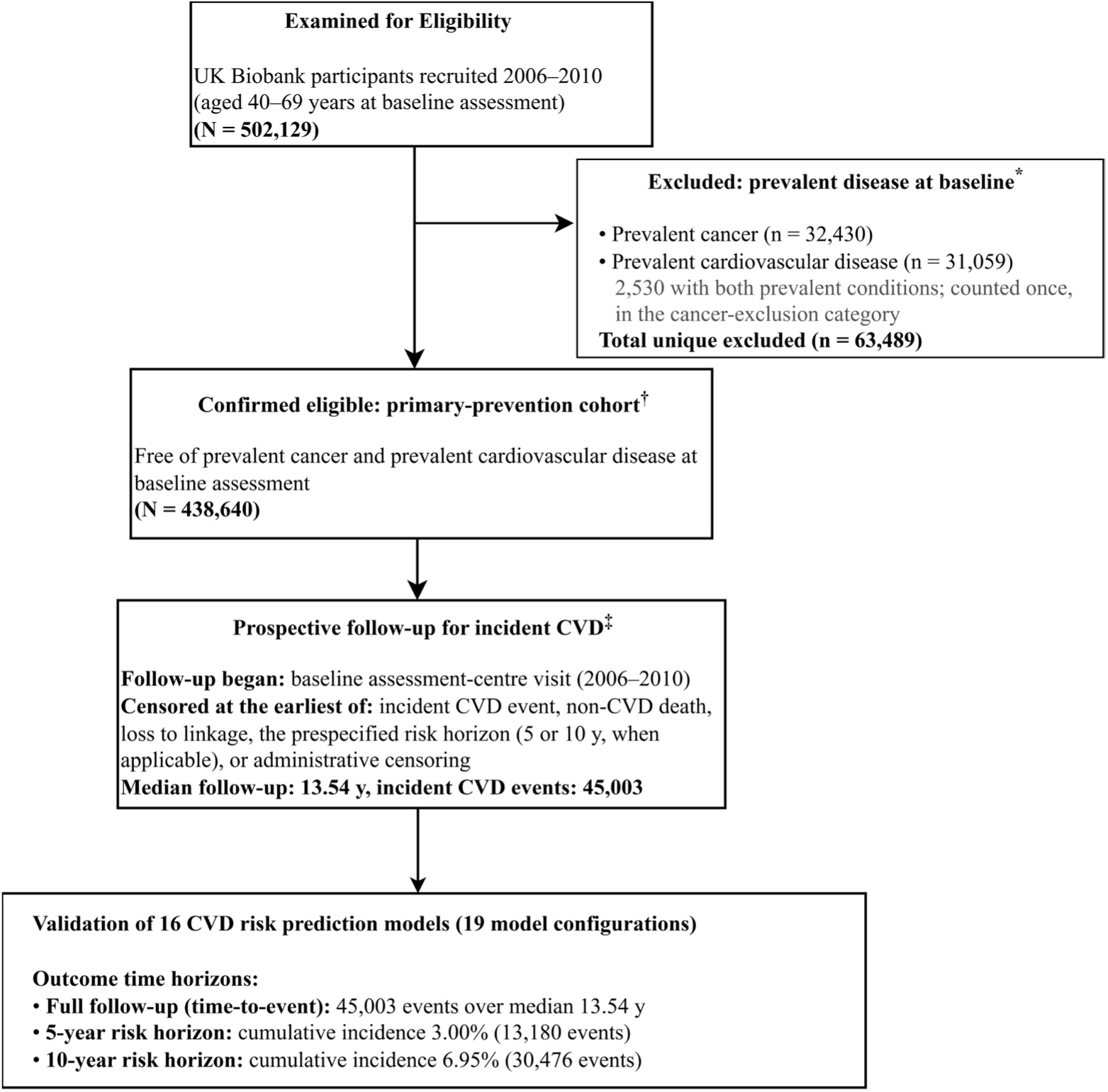
Performance was evaluated on the same analytic sample across three times horizons, full follow-up, 5-year, and 10-year. *Prevalent-disease exclusion categories are non-overlapping by construction: 2,530 participants meeting both criteria were assigned once, under prevalent cancer. Diagnostic codes in Methods. †Full baseline characteristics in Table 1. Predictor missingness was handled by multiple imputation by chained equations (MICE, m = 5); risks and performance metrics were computed per imputed dataset and pooled. ‡Endpoint definition and censoring rule in Methods §“Outcome definition and follow-up“; full ICD-10 code list in Supplementary Table 2.

**Table 1.**
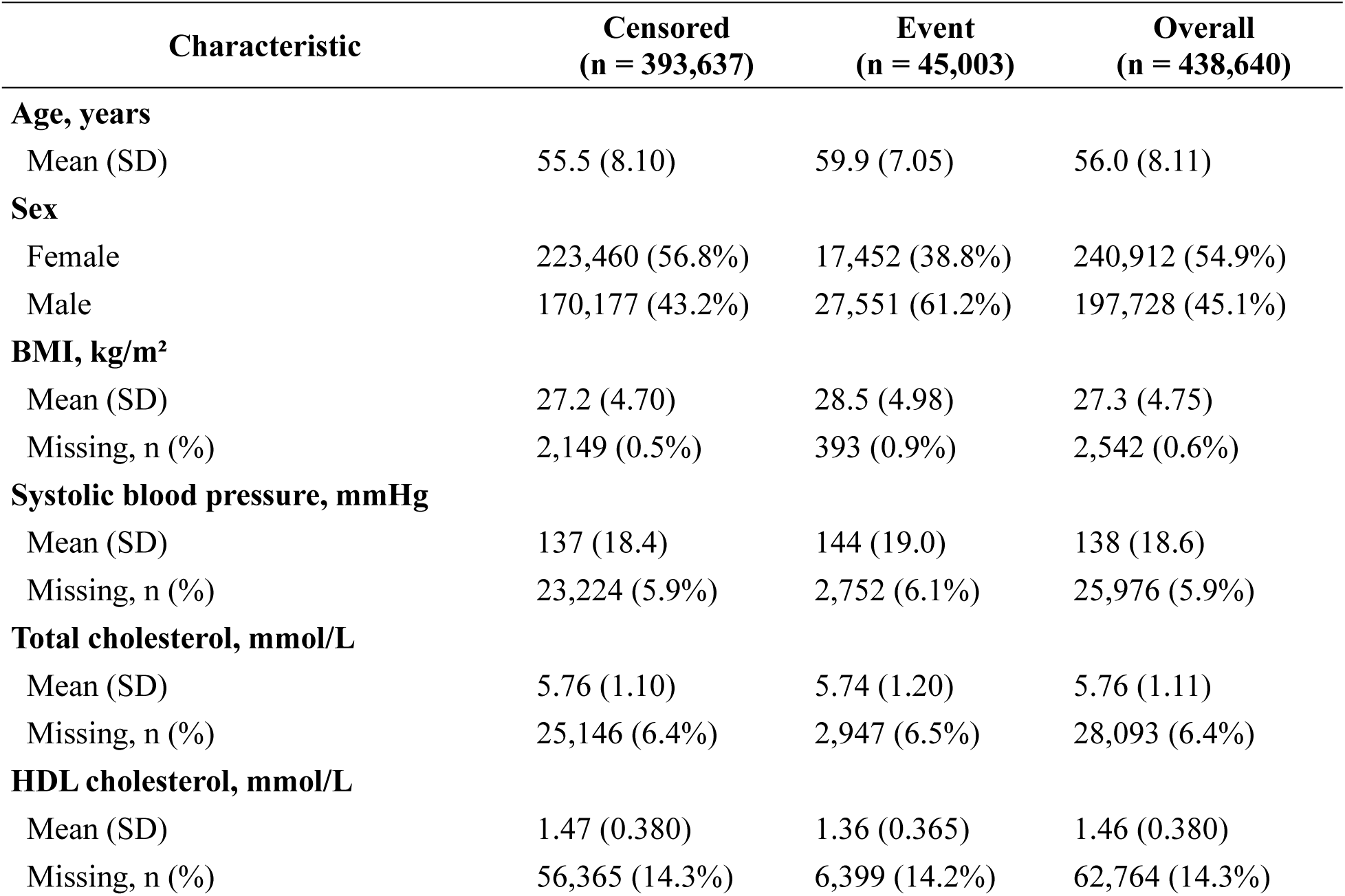
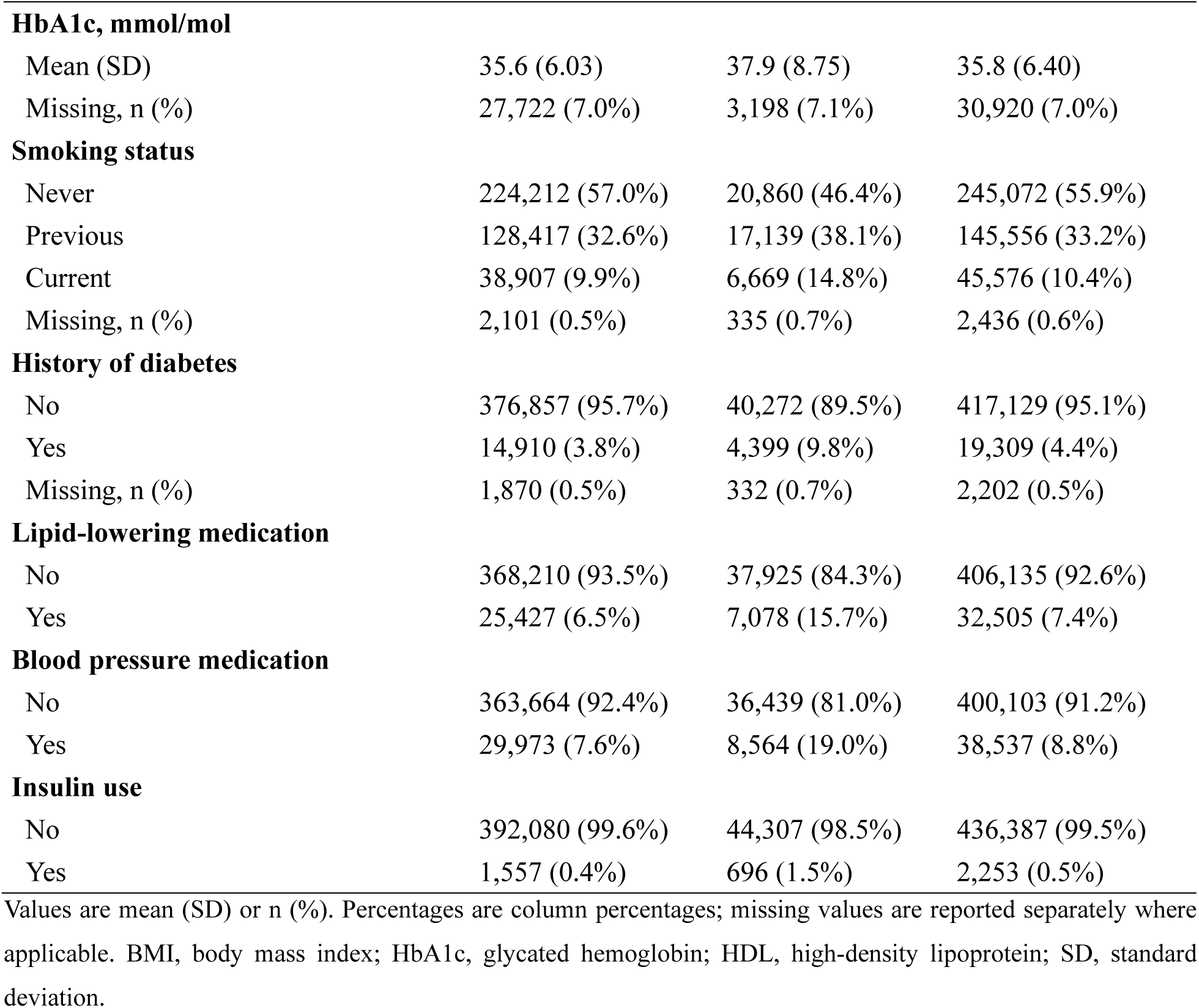
Baseline characteristics of the primary prevention cohort.

### Selection and implementation of risk prediction models

We identified candidate models developed for primary prevention of cardiovascular disease (CVD) through a systematic search of PubMed and major clinical guidelines up to 31 December 2025. Search terms combined specific model names (e.g., “Framingham,” “SCORE,”^28^ “QRISK,” “PREVENT”) with keywords related to risk assessment and prevention (e.g., “absolute risk,” “primary prevention,” “risk score”). Models were required to meet all of the following criteria: (i) were derived from general population cohorts for primary prevention using regression methods (Cox or logistic), (ii) provided full model specifications (coefficients, baseline survival/hazard, and intercept) to enable independent calculation of absolute risk, and (iii) have been recommended by current clinical guidelines or widely cited in practice. Models were also excluded if their intended outcome (e.g., single-endpoint stroke) was incompatible with a composite CVD endpoint.

For each included model, we implemented the risk equations exactly as published, incorporating all non-linear transformations and interaction terms. Where multiple versions of a model existed (e.g., region-specific recalibrations), we selected the version most appropriate for a UK-based or European population. Predictors were mapped to UK Biobank data fields with measurement units, categorical encodings, and reference levels harmonized to the original derivation specifications. Full model details, including derivation cohorts, outcome definitions, and predictor sets, are provided in Supplementary Table 1.

### Outcome definition and follow-up

The primary outcome was a harmonized composite incident CVD endpoint, defined as the first occurrence of ischemic heart disease, sudden cardiac death/cardiac arrest, or stroke, identified using ICD-10 codes from linked hospital inpatient and death registry data (full code list in Supplementary Table S2).

The included risk scores were originally developed for related but non-identical cardiovascular endpoints, including coronary events, ASCVD, fatal CVD, fatal and non-fatal CVD, and broader cardiovascular outcomes. We therefore used a harmonized composite incident CVD endpoint for all models to enable direct comparison within the same primary-prevention cohort. This analysis was not designed to reproduce each model’s native derivation endpoint in isolation. Instead, it addressed a pragmatic public-health question: when alternative risk scores are applied to the same initially disease-free population, how well do they stratify individuals for the subsequent burden of clinically important and potentially preventable cardiovascular events? Applying the same endpoint across models provides a common basis for comparing discrimination, calibration, and clinical utility, while recognizing that differences in calibration may partly reflect endpoint, population, and baseline-risk transportability. Original model-specific endpoints are summarized in Supplementary Table S1.

Where a participant experienced more than one qualifying event, only the first event was retained. Follow-up began at the assessment center visit and ended at the earliest of the primary outcome, death from non-CVD causes, loss to linkage, the prespecified risk horizon, or the administrative censoring date.

### Reporting-quality assessment

We assessed the original development or update publication for each of the 16 models against the 27-item, 52-sub-item TRIPOD+AI (2024) checklist^24^. Each sub-item was scored as fully reported, partially reported, or not reported by two independent raters (Y.H. and S.H.). To enable functional interpretation of reporting patterns, we additionally grouped the 52 sub-items into four conceptual domains: methodological rigor (items 8a–8c, 9a–9c, 10, 11, 13), reproducibility and transparency (items 1, 4, 12g, 18c–18f, 22), clinical implementability (items 3a–3b, 15, 27a–27b), and fairness and equity (items 3c, 14, 19, with cross-cutting items 8a, 9c, 20b, 23a).

### Statistical analysis

**Missing data.** Baseline predictor missingness was handled using Multiple Imputation by Chained Equations (MICE) with five imputed datasets. The imputation models included all baseline predictors entering any of the evaluated risk scores, along with the event indicator and the follow-up time to preserve the associations between predictors and the outcome^29,30^. Continuous variables were imputed using predictive mean matching and categorical variables using logistic regression. Participants with missing values for the primary outcome or follow-up time were excluded prior to imputation. Predicted risks and all downstream performance metrics were computed within each imputed dataset and combined <u>into one</u>.

**Discrimination.** We assessed discrimination at 5- and 10-year horizons using the time-dependent area receiver operating characteristic curve (AUC) estimated via inverse probability of censoring weighting (IPCW) to address right-censoring. Harrell’s C-index was additionally computed, and the C-index between models were estimated with 95% confidence intervals using bootstrap resampling (500 iterations).

**Calibration.** Calibration was assessed using three complementary approaches. First, decile-based calibration plots compared mean predicted risk against KM estimated observed risk within each decile of predicted risk. Second, calibration slope (β) and intercept (α) were estimated under a logistic recalibration framework and indicate perfect calibration. Third, overall prediction error was quantified by the IPCW Brier score and by the Index of Prediction Accuracy (IPA = 1 − Brier_model / Brier_null), where IPA = 1 indicates perfect prediction and IPA = 0 indicates equivalence to a null model.

**Clinical utility.** Net benefit was estimated using decision curve analysis across intervention thresholds from 1% to 30%, with standardized net benefit reported at the four prespecified guideline-anchored thresholds of low (<5%), borderline (5%-7.5%), intermediate (7.5%-10%), and high (>10%) predicted risk^3,31^. Treat-all and treat-none strategies served as reference comparators.

**Integrative performance evaluation.** To synthesize performance across the four domains, we constructed a composite performance assessment (CPA) index. For each of the 19 configurations, four domain-level scores — 10-year time-dependent AUC, Index of Prediction Accuracy, standardized net benefit at the 7.5% intervention threshold, and TRIPOD+AI overall completion rate — were converted to z-scores across models, oriented so that higher values indicated better performance, and averaged with equal weights. Ward’s-linkage hierarchical clustering was applied to the 19 × 4 standardized matrix to identify groups of models with similar performance patterns, with the optimal cluster number selected by joint inspection of silhouette score, gap statistic, and dendrogram cut height. Bootstrap 95% confidence intervals for both the CPA index and cluster assignment were obtained from 500 resamples of the cohort.

**Software.** All analyses were conducted in R (version 4.3). Time-to-event analyses used the survival package; IPCW-based time-dependent AUC and Brier scores used *timeROC* and *riskRegression*; multiple imputation used *mice*; decision curve analysis used *dcurves*; hierarchical clustering used *cluster* and *factoextra*; and visualization used *ggplot2*, *ComplexHeatmap*, and *forestplot*. Full analysis code, predictor-mapping tables, and figure-generation scripts are available at https://github.com/Biomedical-Statistical-Learning/CVD-Scoring-System-Evaluation.

## Results

### Study population and event rates

Of 502,129 UK Biobank participants, we excluded 32,430 participants with prevalent cancer (except for skin cancer), 31,059 with prevalent cardiovascular diseases, leaving 438,640 individuals in the primary analysis cohort (Figure 1). The median age of participants was 57 (IQR = 14) and 240,912 were women (54.90%). 93.80% were of White European ethnicity, 1.97% South Asian, 1.67% Black, and 1.88% other or mixed. <u>Over a median</u> follow-up of 13.54 years, 45,003 incident CVD events were recorded, corresponding to cumulative event rates of 3.00% (n = 13,180) at 5 years and 6.95% (n = 30,476) at 10 years. Baseline characteristics stratified by event status are shown in Table 1. The proportion of missing data across predictor variables was low (range 14.78% to 0.41%; Supplementary Figure 1).

### Reporting quality of original model development studies

We assessed the 17 original development publications against the TRIPOD+AI (2024) checklist, scoring each of the 52 sub-items as reported, not reported, or not applicable (Figure 2). Across all 884 model × item evaluation units, the proportions classified as Yes, No, and Not applicable were 62.3%, 23.2%, and 14.5%, respectively. Across the 756 applicable evaluation units, the overall reporting rate was 72.9%.

**Fig. 2:**
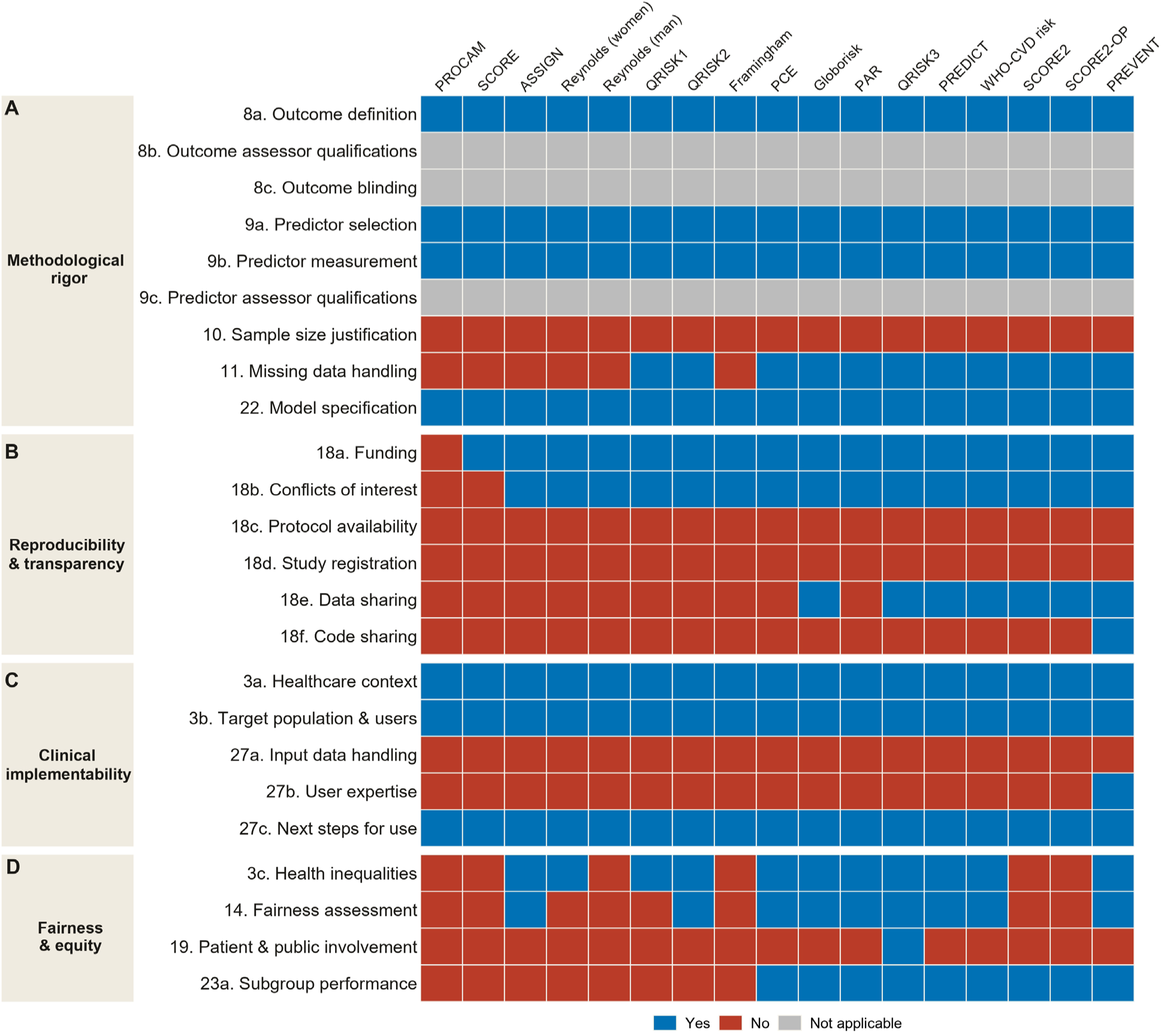
24 TRIPOD +AI subitems selected from the four reporting domains and the full 52-item audit is given in Supplementary. Figure 2. Columns are the 16 models (17 source publications, since Reynolds has separate articles for men and women), ordered chronologically by year of derivation. Rows are TRIPOD+AI sub-items, grouped into four reporting domains.

To characterize reporting patterns beyond overall completion rates, we grouped the 52 items into four functional quality domains: methodological rigor, reproducibility and transparency, clinical implementability, and fairness (details item-to-domain mapping in Supplementary Table 3).

**Methodological rigor** was the best-reported domain. Outcome and predictor definitions (items 8a, 9a–9b) and full model specification (item 22) were fully reported in 100% of models. However, two rigor-related items lagged behind: sample size justification (item 10) was fully reported in none of the models^32^, and missing data handling (item 11) was fully reported in only 64.7% of models. Items related to assessor qualifications or blinding of outcome or predictor assessment (items 8b–8c, 9c) were generally not applicable to these models and therefore rarely contributed to the applicable denominator.

**Reproducibility and transparency** showed the steepest temporal gradient. Among models developed before 2005, protocol availability (item 18c), study registration (item 18d), data sharing (item 18e), and code availability (item 18f) were uniformly unreported. Among models developed after 2015, protocol availability and study registration remained uniformly unreported, whereas data sharing was reported in 85.7% of models and code availability in only 14.3%. Full model specifications enabling independent risk calculation (item 22) were available for 100% of models overall.

**Clinical implementability** was poorly addressed across all eras. Items on clinical context and intended users (items 3a–3b) were universally well reported (100% “Yes”), but guidance on handling missing or poor-quality input data at the point of care (item 27a) and statements on required user expertise (item 27b) were reported in 0% and 5.9% of models, respectively.

**Fairness and equity** was the weakest area overall. A dedicated fairness assessment (item 14) was fully addressed in only 52.9% of models. Patient or public involvement in model design, conduct, or interpretation (item 19) was almost never reported, appearing in only 5.9% of models. Reporting of known health inequalities in the target population (item 3c) and subgroup-specific performance (item 23a) were fully addressed in 64.7% and 52.9% of models, respectively. Even among the most recent and best-reported models, coverage of fairness-related items remained inconsistent (Figure 2).

### Discrimination

Discrimination varied moderately across the 16 prediction models, corresponding to 19 evaluated model configurations. The 10-year time-dependent AUC ranged from 0.668 for QRISK3 to 0.734 for PREDICT, and Harrell’s C-index over 10-year follow-up showed a consistent range of 0.655–0.717 (Figure 3A and 3B. The gap in C-index between the best-performing model (PREDICT) and the lowest-performing model (QRISK3) was 0.062. Five-year AUCs broadly followed the same pattern, ranging from 0.673 to 0.739 (Supplementary Figure 3).

**Fig. 3:**
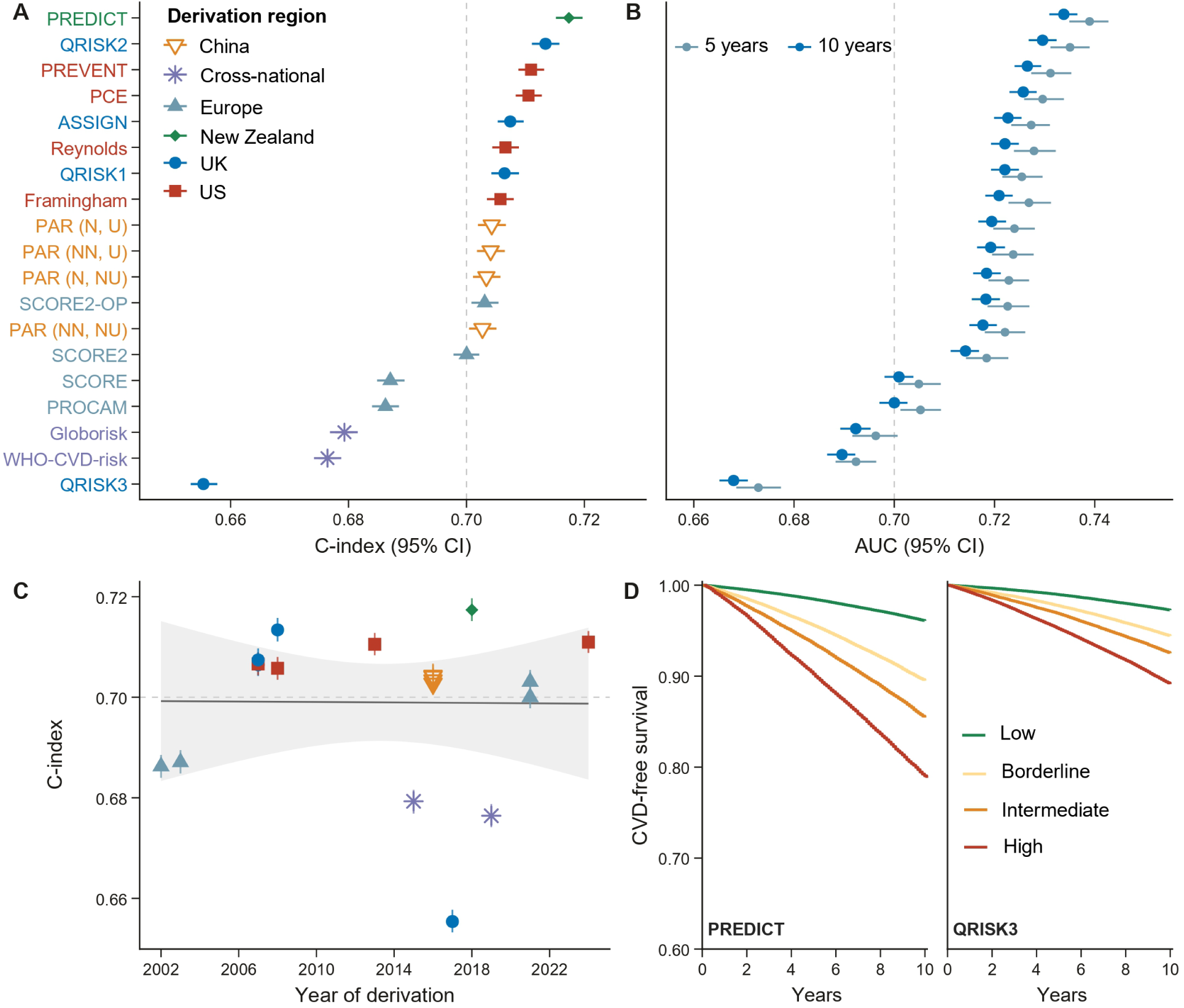
Discrimination of 19 prediction model configurations over 5 and 10 years of follow-up. (A) Harrell’s C-index over 10-year follow-up for each model; models are ordered top-to-bottom by descending C-index. (B) Time-dependent IPCW-AUC at 5-year (light blue) and 10-year (dark blue) horizons with 95% CIs, shown in the same model order as panel A. (C) Year of derivation against Harrell’s C-index, with a linear-regression line and 95% confidence band (slope −2.3 × 10⁻⁵ per year; P = 0.97). (D) Kaplan–Meier CVD-free survival curves over 10-year follow-up for the best-discriminating model (PREDICT, C = 0.717) and the worst (QRISK3, C = 0.655), stratified by predicted 10-year risk category (<5%, 5 to <7.5%, 7.5 to <10%, ≥10%).

Discrimination patterns reflected the geographic context in which models were derived. Scores developed for specific national or regional populations generally clustered in the upper portion of the performance ranking, including UK-derived QRISK2 (C-index 0.713), European SCORE2 (0.700) and SCORE2-OP (0.703), US-derived PREVENT (0.711) and PCE (0.710), New Zealand’s PREDICT (0.717), and the four China-PAR sub-models (range 0.703–0.704). By contrast, the two models designed for cross-national application, Globorisk (C-index 0.679) and WHO-CVD risk (0.676), occupied the lower end of the ranking. QRISK3 (2017), though UK-developed, performed below the other UK-derived models (C-index 0.655), indicating that regional origin alone did not guarantee high discrimination.

A clean temporal gradient was not observed. Among models developed or updated after 2015, performance still spanned a wide range—from the high-discrimination tier (PREDICT (2018), SCORE2 (2021), SCORE2-OP (2021), and PREVENT (2024), all with C-index ≥ 0.700) to lower-performing models such as WHO-CVD risk (2019, C-index 0.676) and QRISK3 (2017, C-index 0.655). Earlier-generation models likewise included both competitive performers (QRISK2 (2008, C-index = 0.713), PCE (2013, C-index = 0.710)) and lower-performing scores (SCORE (2003, C-index = 0.687), PROCAM (2002, C-index = 0.686)). Era of development was therefore less predictive of discrimination than the regional relevance of the derivation population.

### Calibration

A model that ranks risk well may still misestimate its absolute magnitude. To assess this, we compared predicted 10-year absolute risk with observed event rates using decile-based calibration plots, calibration slopes and intercepts, and IPCW Brier scores (Figure 4; Supplementary Figure 4).

**Fig 4:**
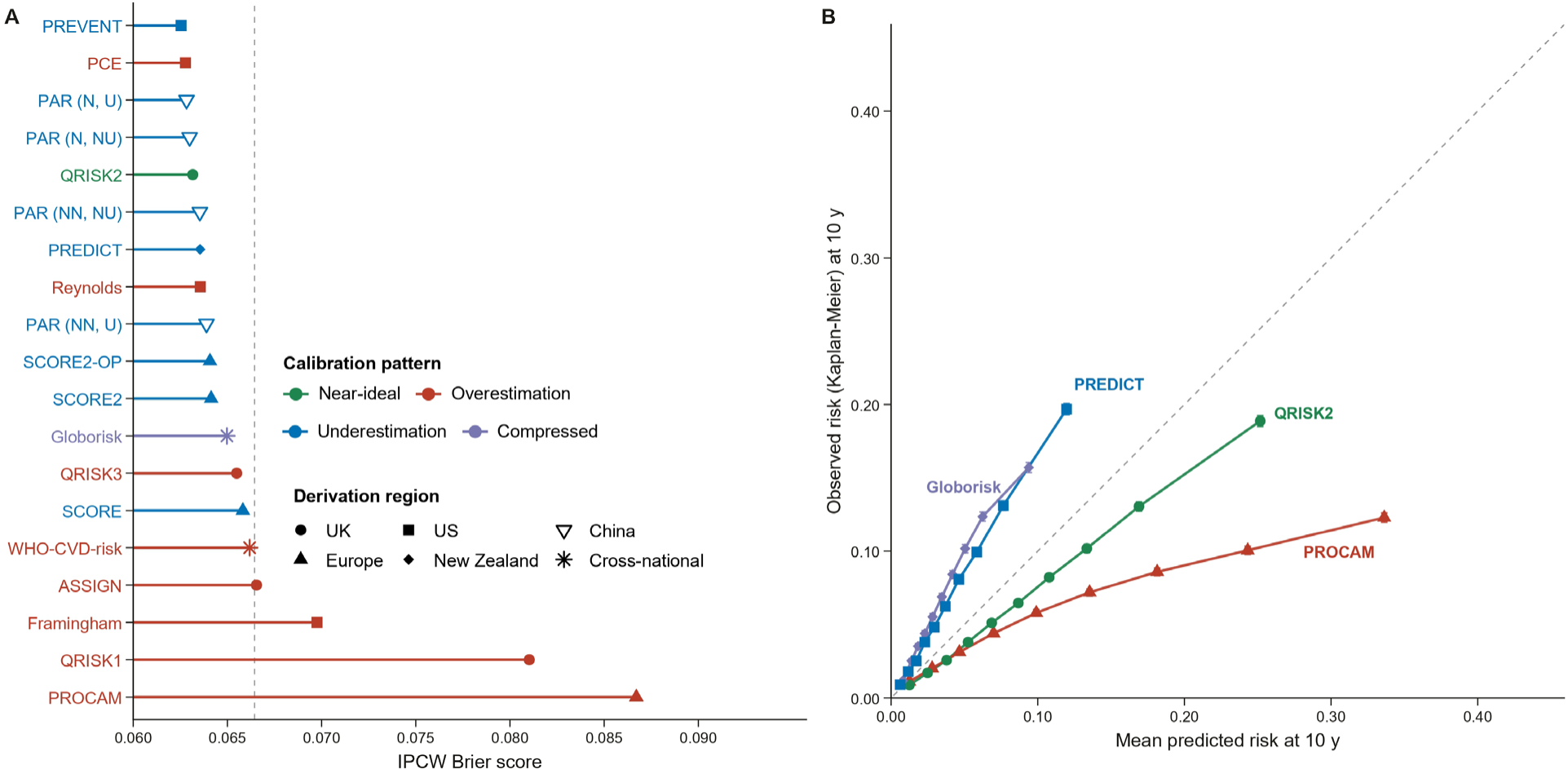
Calibration of predicted versus observed 10-year CVD risk across 19 prediction model configurations. (A) IPCW Brier score for each model, ranked top-to-bottom from lowest (best calibration accuracy) to highest. The vertical dashed line marks the event-rate-implied uninformative-prediction Brier baseline (0.0664). Point color denotes the calibration pattern observed in the manuscript and the shape denotes the derivation region. (B) Calibration curves for four archetype models overlaid on a common set of axes: QRISK2 (near-ideal), PROCAM (overestimation), PREDICT (underestimation), and Globorisk (compressed predicted-risk range). Each point is the mean predicted risk plotted against the KM observed event rate at 10 years within one of 10 quantile groups of predicted risk. Models whose curves lie below y = x over-predict risk relative to the observed event rate, and models whose curves lie above y = x under-predict.

Overall prediction error was modest for most models, although two models stood out as clear outliers. IPCW Brier scores ranged from 0.0625 for PREVENT to 0.0867 for PROCAM (delta = 0.0242), while most of the remaining models clustered within a narrower range of 0.0628-0.0698. Calibration slopes ranged from 0.53 to 1.26 and intercepts from −1.67 to 1.26, indicating that models with broadly similar aggregate accuracy could still diverge materially from observed risk at specific deciles.

Eight models showed clear overestimation, with predicted risks tending to exceed observed event rates, particularly in the higher deciles. The clearest overestimation was seen for PROCAM (slope 0.53; highest-decile predicted 0.33 vs. observed 0.11), with substantial overestimation also evident for QRISK1 (0.43 vs. 0.18), Framingham (0.37 vs. 0.18), ASSIGN (0.31 vs. 0.17), QRISK3 (0.25 vs. 0.14), Reynolds (0.27 vs. 0.19), PCE (0.26 vs. 0.19), and WHO-CVD at higher deciles.

Nine models showed underestimation to varying degrees, with observed event rates generally exceeding predictions. These included PREVENT (slope 1.11; highest-decile predicted 0.15 vs. observed 0.19), PREDICT (0.13 vs. 0.20), SCORE2, SCORE2-OP, SCORE, and the four China-PAR sub-models, although the north-urban and north-nonurban variants lay closer to the reference? line than the two non-north variants. Their predicted risk ranges were also generally narrower, with highest-decile predicted risks typically peaking at 0.08-0.17. QRISK2 was closest to ideal calibration overall, lying near the 45° line across most deciles.

Globorisk showed a third, distinct pattern: predicted values were compressed within a narrow band (0.01-0.09) with limited separation across deciles, even though observed risk rose to 0.15 in the highest group. This compression paralleled its weaker discrimination and would be less amenable to simple linear recalibration of the slope and intercept alone.

Among the China-PAR sub-models, the north-urban variant came closest to ideal calibration, with north-nonurban also performing relatively well, whereas the two non-north variants were somewhat less well calibrated. PREVENT achieved the lowest overall error despite mild underestimation, whereas PROCAM had the highest. Most models would therefore require population-specific recalibration before clinical application in a UK primary prevention setting. At the shorter time horizon, patterns were broadly similar.

### Clinical utility

The preceding analyses showed that models differed in both discrimination and calibration; we next evaluated whether these differences translated into more clinically useful treatment decisions. Specifically, we compared standardized net benefit across intervention thresholds from 5% to 20%, using treat-all and treat-none as reference strategies (Figure 5; Supplementary Figure 5).

**Fig 5:**
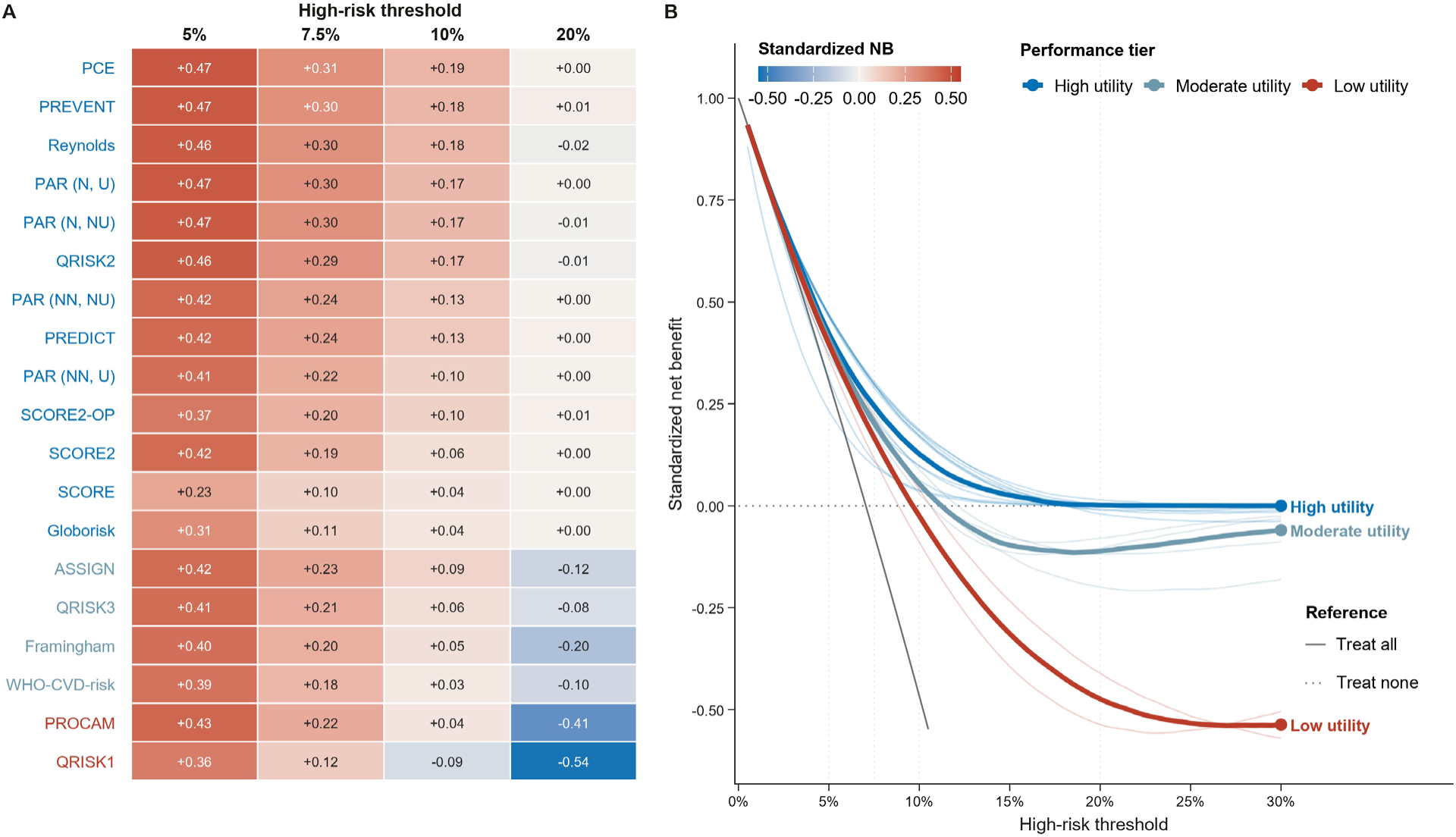
Clinical utilities at a 10-year horizon. (A) Heatmap of standardized net benefit (NB) at the four manuscript-relevant high-risk thresholds (5%, 7.5%, 10%, and 20%). Rows are ordered top-to-bottom by descending NB at the 10% threshold, and each cell is labelled with its numeric NB. Tier assignment was derived from Ward hierarchical clustering (k = 3) of the per-model NB curves over the full threshold grid, and the Y-axis labels are colored by performance tier: high utility (blue, n = 13), moderate utility (steel, n = 4), and low utility (red, n = 2). (B) Decision-curve analysis across high-risk thresholds 0.5–30%. Thick lines show the per-tier median NB trajectory (high, moderate, low utility), and faint colored lines behind them are the individual model curves within each tier, drawn with equal visual weight. Treat-all (solid grey) and treat-none (dotted grey) are shown as reference strategies.

At thresholds of 5% and 7.5%, all models yielded positive net benefit, indicating that each score outperformed blanket strategies within this lower-threshold interval. At 10%, however, one model (QRISK1) fell below the treat-none baseline, whereas the remaining models still showed positive net benefit. PREVENT and PCE consistently ranked among the top-performing models: PREVENT achieved the highest standardized net benefit at 5% (0.467), whereas PCE was highest at 7.5% (0.305) and 10% (0.187).

Other strong performers in this range included Reynolds (0.463, 0.296, and 0.181 at 5%, 7.5%, and 10%, respectively), QRISK2 (0.462, 0.288, 0.168), and the north urban and north nonurban China-PAR sub-models. By contrast, SCORE showed the lowest net benefit at 5% and 7.5%, while Globorisk was among the lowest positive models at 10%; QRISK1 was the only model with negative net benefit at this threshold.

Above 10%, models diverged more sharply. At a threshold of 20%, net benefit for most models approached zero, and around half were at or below zero. QRISK1 showed the most pronounced negative value (−0.53), followed by PROCAM (−0.41) and Framingham (−0.20). PREVENT retained the clearest positive net benefit (0.014), while several other models, including PREDICT, SCORE, SCORE2-OP, and selected China-PAR sub-models, remained only marginally above zero; PCE, Globorisk, and SCORE2 were effectively neutral at this threshold. These patterns suggest that models combining competitive discrimination with better calibration deliver more robust decision utility when treatment thresholds become more stringent.

### Composite performance assessment identified a recommended cluster of models

Discrimination, calibration, clinical utility, and reporting quality each captured different aspects of model suitability. To determine whether these dimensions converged on a coherent ranking, we integrated them into a composite performance assessment (CPA) index combining 10-year AUC, IPA, standardized net benefit at the 7.5% threshold, and TRIPOD+AI completion (Figure 6).

**Fig 6:**
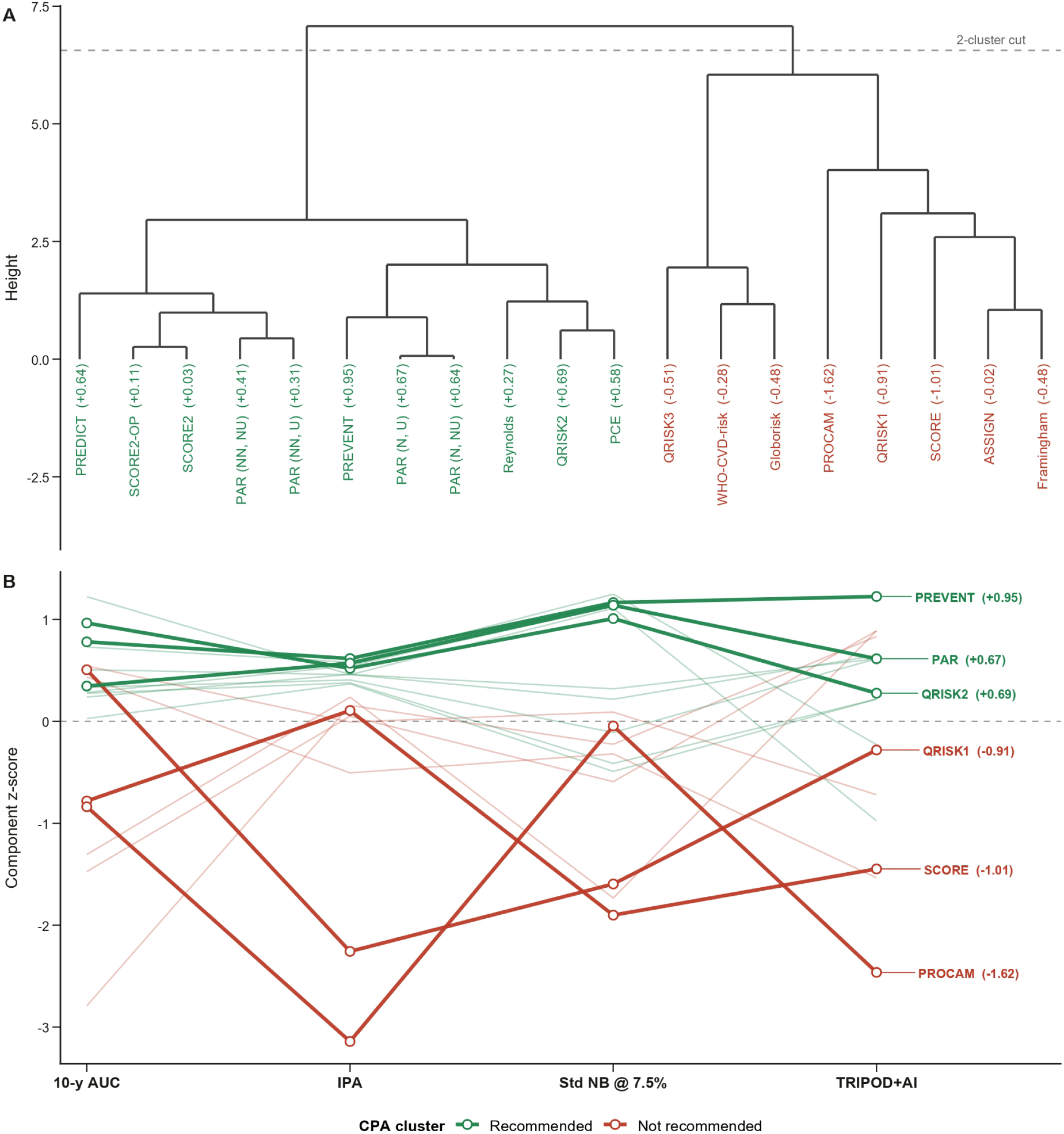
Composite performance assessment integrating discrimination, calibration, clinical utility, and reporting quality. (A) Hierarchical-clustering dendrogram of all 19 configurations, computed on Euclidean distances between their four CPA component z-scores with Ward linkage. The horizontal dashed line marks the two-cluster cut that separates the recommended cluster (n = 11, green) from the not-recommended cluster (n = 8, red). CPA index values range from +0.95 (PREVENT, highest) to −1.62 (PROCAM, lowest). (B) Parallel-coordinates plot of each model’s z-score profile across the four CPA components (10-year AUC, IPA, standardized NB at 7.5%, TRIPOD+AI). All 19 configurations are drawn as faint lines; the three highest-ranked models (PREVENT, QRISK2, PAR (N, U)) and the three lowest-ranked models (QRISK1, SCORE, PROCAM) are highlighted with thicker lines and direct labels reporting each model’s CPA index.

Hierarchical clustering separated the 19 evaluated model configurations into two groups: a recommended cluster containing 11 models and an unrecommended cluster of 8. CPA index values ranged from 0.95 (95% CI 0.93–0.97) for PREVENT to −1.62 (95% CI −1.65 to −1.59) for PROCAM. he recommended cluster comprised PREVENT (0.95), QRISK2 (0.69), China-PAR–north-urban (0.67), China-PAR–north-non-urban (0.64), PREDICT (0.64), PCE (0.58), and SCORE2, SCORE2-OP, Reynolds, and the two southern China-PAR sub-models. The unrecommended cluster comprised SCORE, QRISK1, QRISK3, Globorisk, WHO-CVD risk, Framingham, ASSIGN, and PROCAM (Supplementary Figure 6A).

The heatmap (Supplementary Figure 6B) showed that this separation was driven primarily by differences in multidimensional balance rather than by any single component alone. PREVENT displayed the most consistently favorable profile, with above-average z-scores across all four dimensions, indicating broadly strong performance. QRISK2 and the two northern China-PAR sub-models showed a similar pattern, combining favorable discrimination, IPA, and DCA with acceptable reporting completeness. PREDICT was driven primarily by its strong discrimination signal, with a more modest contribution from clinical utility. PCE remained in the recommended cluster despite weaker TRIPOD component, indicating that stronger discrimination and decision performance compensated for its lower TRIPOD+AI score. SCORE2 and SCORE2-OP also remained on the recommended side, though their composite advantage was smaller than that of the highest-ranked scores.

Models in the unrecommended cluster were generally showed deficits across multiple domains. QRISK3 and WHO-CVD risk retained acceptable TRIPOD-related profiles but were pulled downward by weaker discrimination and clinical utility. Globorisk was unfavorable across both discrimination and clinical utility. PROCAM and QRISK1 were penalized most strongly, with markedly low composite scores driven largely by poor IPA-related performance and limited overall balance across domains. Framingham and ASSIGN occupied a less extreme but still unrecommended position, with moderate performance in isolated dimensions insufficient to offset broader multidimensional weaknesses. The CPA framework therefore identifies PREVENT, QRISK2, the northern China-PAR variants, PREDICT, and PCE as the strongest candidate models in the UK Biobank context.

## Discussion

In this head-to-head external validation study, we compared 16 widely used cardiovascular risk-prediction models, corresponding to 19 model configurations, in 438,640 UK Biobank participants without prevalent cardiovascular disease or cancer at baseline. Over a median follow-up of 13.5 years, 45,003 incident cardiovascular events occurred. Four main findings emerged. First, discriminatory performance varied across models, however, newer models did not uniformly outperform older scores. Second, calibration varied more widely than discrimination, with several models substantially overestimating or underestimating absolute risk. Third, these differences affected clinical utility^33^, particularly at higher treatment thresholds. Finally, reporting quality was uneven, especially for implementation guidance, transparency, sample-size justification, and fairness-related reporting. Overall, PREVENT, QRISK2, PREDICT, PCE, and the northern China-PAR variants showed the most favorable multi-domain profiles in this cohort.

These findings show that cardiovascular risk scores should not be treated as interchangeable tools. Risk models are shaped by their derivation populations, outcome definitions, baseline event rates, predictor definitions, and healthcare systems. When transferred to a new setting, even established guideline-endorsed models may perform differently. This matters because absolute risk estimates are used to guide statin therapy, blood pressure treatment, lifestyle counselling, and shared decision-making^3,6^. A model that ranks individuals reasonably well may still lead to inappropriate treatment decisions if it misestimates absolute risk.

Discrimination varied across models, though most contemporary scores performed within a moderate range^34^. PREDICT had the highest discrimination, while QRISK3, WHO-CVD risk, and Globorisk performed less well. Models developed in specific national or regional populations generally performed better than models designed for broad global use. However, regional origin alone was not sufficient. QRISK3 was developed in the UK but showed lower discrimination than QRISK2, whereas US-derived PREVENT and PCE performed competitively. These results suggest that transportability depends not only on geography, but also on endpoint definition, predictor implementation, treatment patterns, and population structure.

Calibration was the most clinically important source of variation^35^. PROCAM, Framingham, ASSIGN^36^, and QRISK1 substantially overestimated risk^37^, particularly among individuals in the highest predicted-risk deciles. Such overestimation could increase unnecessary treatment, monitoring, and patient concern if these models were applied without recalibration. Conversely, PREVENT, PREDICT, SCORE, SCORE2, SCORE2-OP, and the China-PAR sub-models tended to underestimate risk to varying degrees, which could delay preventive treatment in some individuals. QRISK2 was closest to ideal calibration overall, whereas PREVENT had the lowest prediction error despite mild underestimation. These results highlight the need to assess calibration graphically and quantitatively before adopting any risk score in practice.

Decision curve analysis confirmed that statistical differences had clinical consequences^38^. At lower thresholds, all models provided positive net benefit, supporting the general value of structured risk stratification. At higher thresholds, however, performance diverged. PREVENT and PCE retained favorable clinical utility, whereas QRISK1, PROCAM, and Framingham became neutral or potentially harmful. This is important because treatment thresholds determine who is labeled high risk and who receives long-term preventive therapy.

PREVENT showed the most balanced overall performance in this analysis. Its strengths likely reflect its contemporary derivation, large development population, and inclusion of cardiovascular–kidney–metabolic predictors^39^. However, these findings should not be interpreted as evidence that PREVENT should automatically replace existing UK or European scores. UK Biobank is not fully representative of routine primary-care populations, and the harmonized outcome used in this study differs from the original endpoints of some models^40,41^. Rather, PREVENT should be considered a strong candidate for further evaluation, recalibration, and comparison with QRISK and SCORE2-based strategies in representative UK and European healthcare settings^2,4^.

The results also have implications for established UK and European models. QRISK2^42^ performed well, particularly for calibration, supporting the value of models developed in settings whose population structure, predictor definitions, and baseline risk are broadly similar to the evaluation setting. SCORE2 and SCORE2-OP showed reasonable discrimination but tended to underestimate risk, suggesting that further local recalibration may be required when applied outside their intended risk-region context or when outcome definitions differ. The weaker performance of QRISK3 should be interpreted cautiously because some predictors may not have been captured in UK Biobank with the same completeness or definitions as in routine primary-care data^43^.

The reporting-quality assessment adds a further message^24^. Most development studies reported predictors, outcomes, and model equations sufficiently for implementation. However, important gaps remained. Sample size justification was rarely provided, practical guidance for handling missing or poor-quality inputs was largely absent, and data/code transparency was inconsistent. Fairness reporting was particularly limited, including subgroup performance, health inequality considerations, and patient or public involvement. These gaps are important because risk scores increasingly influence access to preventive treatment.

In this study, we provide a comprehensive comparison of widely used cardiovascular risk scores in a single large cohort, using harmonized predictor mapping, outcome ascertainment, follow-up definitions, and statistical methods. The large number of events enabled stable assessment at 5- and 10-year horizons. We evaluated discrimination, calibration, prediction error, clinical utility, and reporting quality, rather than relying on a single performance metric.

In conclusion, cardiovascular risk prediction models differ substantially when applied to a single large cohort. No single metric is sufficient to judge model suitability, and no model should be assumed transportable without external validation^44^. In UK Biobank, PREVENT showed the most balanced overall profile, while QRISK2, PREDICT, PCE, and selected China-PAR variants also performed favorably. These findings support a more rigorous approach to risk score selection: external validation, local recalibration, decision curve analysis, transparent reporting, and fairness assessment should be prerequisites for implementation in cardiovascular primary prevention.

## Data Access and Code Availability

Data were obtained from UK Biobank under approved application 160957. Individual-level UK Biobank data cannot be shared by the authors but are available to eligible researchers through the UK Biobank Access Management System. Analysis code, including model implementation, predictor harmonization, performance evaluation, and figure-generation scripts, is available at: https://github.com/Biomedical-Statistical-Learning/DANN_cell_like_me.

## Acknowledgements

The authors thank Biao Xie for their assistance with data acquisition. J.Z. was supported by the Chongqing Municipal Science and Technology Bureau (Grant No. CSTB2024YCJH-KYXM0098) and the Venture and Innovation Support Program for Chongqing Overseas Returnees (Grant No. 203011220240002).

## Author Contributions

Y.H. and J.Z. conceived the study and drafted the manuscript. Y.H., S.H., J.W., and J.Z. performed the data analysis. Z.D., J.W., Z.Z., P.J., H.H., T.L., and J.Z. contributed to interpretation of the findings and critical revision of the manuscript. H.H., T.L., and J.Z., provided methodological guidance and supervision. All authors reviewed and approved the final manuscript.

## References

1. Di Cesare, M. et al. The Heart of the World. Glob. Heart 19, 11 (2024).

2. Hippisley-Cox, J., Coupland, C. & Brindle, P. Development and validation of QRISK3 risk prediction algorithms to estimate future risk of cardiovascular disease: prospective cohort study. BMJ 357, j2099 (2017).

3. Arnett, D. K. et al. 2019 ACC/AHA guideline on the primary prevention of cardiovascular disease: a report of the American College of Cardiology/American Heart Association Task Force on Clinical Practice Guidelines. Circulation 140, e596–e646 (2019).

4. SCORE2 working group and ESC Cardiovascular risk collaboration. SCORE2 risk prediction algorithms: new models to estimate 10-year risk of cardiovascular disease in Europe. Eur. Heart J. 42, 2439–2454 (2021).

5. SCORE2-OP working group and ESC Cardiovascular risk collaboration. SCORE2-OP risk prediction algorithms: estimating incident cardiovascular event risk in older persons in four geographical risk regions. Eur. Heart J. 42, 2455–2467 (2021).

6. Visseren, F. L. J. et al. 2021 ESC Guidelines on cardiovascular disease prevention in clinical practice. Eur. Heart J. 42, 3227–3337 (2021).

7. D’Agostino, R. B. Sr., et al. General cardiovascular risk profile for use in primary care: the Framingham Heart Study. Circulation 117, 743–753 (2008).

8. Assmann, G., Cullen, P. & Schulte, H. Simple scoring scheme for calculating the risk of acute coronary events based on the 10-year follow-up of the Prospective Cardiovascular Münster (PROCAM) Study. Circulation 105, 310–315 (2002).

9. Hippisley-Cox, J. et al. Derivation and validation of QRISK, a new cardiovascular disease risk score for the United Kingdom: prospective open cohort study. BMJ 335, 136 (2007).

10. Goff, D. C. Jr., et al. 2013 ACC/AHA guideline on the assessment of cardiovascular risk. Circulation 129, S49–S73 (2014).

11. Yang, X. et al. Predicting the 10-year risks of atherosclerotic cardiovascular disease in Chinese population: the China-PAR Project. Circulation 134, 1430–1440 (2016).

12. Pylypchuk, R. et al. Cardiovascular disease risk prediction equations in 400 000 primary care patients in New Zealand: a derivation and validation study. Lancet 391, 1897–1907 (2018).

13. Ridker, P. M., Buring, J. E., Rifai, N. & Cook, N. R. Development and validation of improved algorithms for the assessment of global cardiovascular risk in women: the Reynolds Risk Score. JAMA 297, 611–619 (2007).

14. Ridker, P. M., Paynter, N. P., Rifai, N., Gaziano, J. M. & Cook, N. R. C-reactive protein and parental history improve global cardiovascular risk prediction: the Reynolds Risk Score for men. Circulation 118, 2243–2251 (2008).

15. Kaptoge, S. et al. World Health Organization cardiovascular disease risk charts: revised models to estimate risk in 21 global regions. Lancet Glob. Health 7, e1332–e1345 (2019).

16. Hajifathalian, K. et al. A novel risk score to predict cardiovascular disease risk in national populations (Globorisk): a pooled analysis of prospective cohorts and health examination surveys. Lancet Diabetes Endocrinol. 3, 339–355 (2015).

17. Khan, S. S. et al. Development and validation of the American Heart Association’s PREVENT equations. Circulation 149, 430–449 (2024).

18. DeFilippis, A. P. et al. An analysis of calibration and discrimination among multiple cardiovascular risk scores in a modern multiethnic cohort. Ann. Intern. Med. 162, 266–275 (2015).

19. Damen, J. A. A. G. et al. Prediction models for cardiovascular disease risk in the general population: systematic review. BMJ 353, i2416 (2016).

20. Parsons, R. E. et al. Independent external validation of the QRISK3 cardiovascular disease risk prediction model using UK Biobank. Heart 109, 1690–1697 (2023).

21. Ambrosio, M. et al. Performance of PREVENT and pooled cohort equations for predicting 10-year ASCVD risk in the UK Biobank. Am. J. Prev. Cardiol. 22, 101009 (2025).

22. Muntner, P. et al. Validation of the atherosclerotic cardiovascular disease Pooled Cohort Risk Equations. JAMA 311, 1406–1415 (2014).

23. Pennells, L. et al. Equalization of four cardiovascular risk algorithms after systematic recalibration: individual-participant meta-analysis of 86 prospective studies. Eur. Heart J. 40, 621–631 (2019).

24. Collins, G. S. et al. TRIPOD+AI statement: updated guidance for reporting clinical prediction models that use regression or machine learning methods. BMJ 385, e078378 (2024).

25. Collins, G. S. et al. Transparent reporting of a multivariable prediction model for individual prognosis or diagnosis (TRIPOD): the TRIPOD statement. Ann. Intern. Med. 162, 55–63 (2015).

26. Riley, R. D. et al. Evaluation of clinical prediction models (part 2): how to undertake an external validation study. BMJ 384, e074820 (2024).

27. Sudlow, C. et al. UK Biobank: an open access resource for identifying the causes of a wide range of complex diseases of middle and old age. PLoS Med. 12, e1001779 (2015).

28. Conroy, R. M. et al. Estimation of ten-year risk of fatal cardiovascular disease in Europe: the SCORE project. Eur. Heart J. 24, 987–1003 (2003).

29. van Buuren, S., Boshuizen, H. C. & Knook, D. L. Multiple imputation of missing blood pressure covariates in survival analysis. Stat. Med. 18, 681–694 (1999).

30. Sterne, J. A. C. et al. Multiple imputation for missing data in epidemiological and clinical research: potential and pitfalls. BMJ 338, b2393 (2009).

31. US Preventive Services Task Force. Statin use for the primary prevention of cardiovascular disease in adults: US Preventive Services Task Force recommendation statement. JAMA 328, 746–753 (2022).

32. Riley, R. D. et al. Calculating the sample size required for developing a clinical prediction model. BMJ 368, m441 (2020).

33. Yadlowsky, S. et al. Clinical implications of revised Pooled Cohort Equations for estimating atherosclerotic cardiovascular disease risk. Ann. Intern. Med. 169, 20–29 (2018).

34. Steyerberg, E. W. & Vergouwe, Y. Towards better clinical prediction models: seven steps for development and an ABCD for validation. Eur. Heart J. 35, 1925–1931 (2014).

35. Van Calster, B. et al. Calibration: the Achilles heel of predictive analytics. BMC Med. 17, 230 (2019).

36. Woodward, M., Brindle, P., Tunstall-Pedoe, H. & SIGN Group on Risk Estimation. Adding social deprivation and family history to cardiovascular risk assessment: the ASSIGN score from the Scottish Heart Health Extended Cohort (SHHEC). Heart 93, 172–176 (2007).

37. Brindle, P. et al. Predictive accuracy of the Framingham coronary risk score in British men: prospective cohort study. BMJ 327, 1267 2003).

38. Vickers, A. J. & Elkin, E. B. Decision curve analysis: a novel method for evaluating prediction models. Med. Decis. Making 26, 565–574 (2006).

39. Khan, S. S. et al. Novel prediction equations for absolute risk assessment of total cardiovascular disease incorporating cardiovascular-kidney-metabolic health: a scientific statement from the American Heart Association. Circulation 148, 1982–2004 (2023).

40. Batty, G. D., Gale, C. R., Kivimäki, M., Deary, I. J. & Bell, S. Comparison of risk factor associations in UK Biobank against representative, general population based studies with conventional response rates: prospective cohort study and individual participant meta-analysis. BMJ 368, m131 (2020).

41. van Alten, S., Domingue, B. W., Faul, J., Galama, T. & Marees, A. T. Reweighting UK Biobank corrects for pervasive selection bias due to volunteering. Int. J. Epidemiol. 53, dyae054 (2024).

42. Hippisley-Cox, J. et al. Predicting cardiovascular risk in England and Wales: prospective derivation and validation of QRISK2. BMJ 336, 1475–1482 (2008).

43. Hippisley-Cox, J. et al. Development and validation of a new algorithm for improved cardiovascular risk prediction. Nat. Med. 30, 1440–1447 (2024).

44. Cook, N. R. Use and misuse of the receiver operating characteristic curve in risk prediction. Circulation 115, 928–935 (2007).

